# Epidemiology and Cluster Predictions on Prevention Strategies of SARS-CoV-2 Cases from 2019-2024 in Selected Districts of Zambia

**DOI:** 10.1101/2025.09.27.25336412

**Authors:** Enock Wantakisha, Stanley Nyirenda, Mony Narayani

**Affiliations:** Department of Public Health, Chreso University, Lusaka, Zambia; Department of Immunology and Pathology, Central Veterinary Research Institute, Lusaka, Zambia; Department of Public Health and Epidemiology, Tropical Disease Research Center, Ndola, Zambia

## Abstract

**Background:** The Severe Acute Respiratory Syndrome Coronavirus 2 (SARS-CoV-2) has continued to be a global health threat despite many interventions put with new variants noticed. There is scarce information on SARS-CoV-2 predisposing factors in relation to the proximate-determinant framework in Zambia. This study aimed to determine epidemiological and cluster predictions on prevention strategies of SARS-CoV-2 cases in Zambia 2019-2024 in rural and urban settings.

**Materials and Methods:** An exploratory sequential mixed method study conducted in three districts; Ndola (Urban), Kafue (Peri-urban) and Lufwanyama (Rural). The target population were all the reported and recorded SARS CoV-2 cases above 15 years and all active cases. Study conducted; 81 random hospital-patient reviews, 21 face-to-face interviews, 5 systematic reviews and an addition 426 telephone survey. The participants were sampled using mixed mode sampling technique. Data was collected using records, structured questionnaires and interviews. STATA(Ver.17) as well as thematic analysis were employed.

**Results:** 624 participated in this study with 528 cases having had tested or confirmed records of SARS CoV-2 test status with mean age 32 (SD10.3). There were more 271 (51.33%) males than 257 (48.7%) females with 80% listing home and work (67%) as places where they had spent over 8 hours of their time respectively. Persons observed at risk of severe cases of infection were; elderly males above 49 years (cOR=2.13, 95% CI: 1.16-3.89, p<0.014), and those who had other co-morbidities (43%). Participants aged 49 years and above and had experienced symptoms of SARS-CoV-2 had a significant effect on testing positive. (p<0.001). Twenty-one (21) were active cases with a pooled prevalence of 3.7%.

**Conclusion:** Majority of the population were knowledgeable about the causes, transmission risks and the SARS CoV-2 prevention guidelines. Failure to adhere to the stipulated SARS CoV-2 prevention guidelines accounted for most of the increases transmission rate among the sub-populations. Adults above 49 years with other co-commodities and not vaccinated were the most affected with the infection

## Introduction

Many Global health diseases have continued to be a challenge in management, control and prevention with many strategies developed (1-3). Such among these is the Severe Acute Respiratory Syndrome Coronavirus 2 (SARS COV-2) infection, a contagious pandemic disease which affected many countries especially starting in 2019 to 2024. Coronavirus disease 2019 is a highly contagious infectious disease caused by SARS-CoV-2 (2, 3, 4), (5, 6). Many countries developed strategies and strengthened their health systems to be able to respond and manage this emerging disease.

SARS CoV-2 infection was first identified in 2019 in Wuhan, the capital of Hubei Province, China (7), and had spread globally, resulting in the 2019–2020 and 2021-2023 global pandemic with the third wave having isolated incidences worldwide reported. Spatial incidences are still being recorded across high-risk populations though its actively monitored. SARS-CoV-2 had infected over 65 million people and caused over 1.6 million deaths, globally (2, 4, 5). Many countries in Africa had expanded their responses to the epidemic; and increased preventive measures, reviewed policies, increased interventions such as access to treatment for hospitalized patients, increased access to vaccines and increased awareness to SARS-CoV-2 prevention methods (8-12) among populations. Despite these efforts the pandemic still presents as a public health threat worldwide with new variants reported in many countries (9, 12, 13).

Coronaviruses are a large family of viruses that can cause illness in animals or humans. There are four main sub-groupings of coronaviruses, known as *alpha, beta, gamma*, and *delta*. In humans there are seven known coronaviruses that cause respiratory infections (11, 14, 15). The seven coronaviruses that can infect humans are; 229E (alpha coronavirus), NL63 (alpha coronavirus), OC43 (beta coronavirus), HKU1 (beta coronavirus), MERS-CoV (Middle East respiratory syndrome), SARS-CoV (severe acute respiratory syndrome) and SARS-CoV-2 (Novel coronavirus that causes coronavirus disease 2019, or COVID-19). The virus is single-stranded RNA continuously evolves as changes in the genetic code (genetic mutations) occur during replication of the genome. Genetic lineages of SARS-CoV-2 have been emerging and routinely monitored through epidemiological investigations (9, 16), virus genetic sequencing-based surveillance and laboratory studies have become important(17, 18). The variant classification scheme defines classes of SARS-CoV-2 variants as; variant being monitored (VBM), Variants of interest (VOI), Variants of concern (VOC) and Variants of High Consequences (VOHC) (5, 8, 9, 19).

The impact of this genetic diversity of SARS-CoV-2 in public health necessitates surveillance strategies that systematically sample and characterize representative and predominant strains from populations whilst concomitantly monitoring proximate indicators and any other factors that may be associated with the infection. This background is critically important whilst estimating the magnitude of the SARS-CoV-2 epidemic (10, 20). SARS-CoV-2 can be transmitted through respiratory droplets or smaller aerosols from mouth or nose, which can spread when an infected person coughs, sneezes, speaks or breathes heavily. Therefore, WHO adopted a one-Meter social distancing policy.

Disease trends provide information about the past and current spread, and may be extremely useful in predicting future patterns of the disease and similar infections (18, 21), which in turn is important in adjusting public health policy and interventions. Establishing a functional surveillance system is the best way to do this (21-23). Globally the magnitude of the SARS-CoV pandemic has been estimated through cross-sectional surveys in risk populations and national population-based surveys. Epidemiological surveillances when carried out correctly, can provide more extensive description and analysis of the trends and determinants of the infection. In Zambia, national demographic surveys were conducted in; 2004, 2007, 2013, 2018 and more recent 2024 to understand the impact of interventions and epidemiology of diseases or conditions on the population except all the surveys never included the transmission, prevention or control related to SARS CoV-2 infection (2, 24). Additionally, very scarce information exists in Zambia on the patterns, trends, transmission rates and determinants of SARS-CoV-2 infection. Such information is critical to foster equity-based resource allocation (18, 25, 26). Majority of research studies available on SARS CoV-2 lacks the correlation of conceptual frameworks with the epidemiology of the disease in the sub-populations. It’s hypothesized that epidermic emergency responses by Ministry of Health-Zambia, National Public Health Institute could be more effective if the patterns, trends, high risk groups on the disease are known.

The focus for this study was thus to determine key distribution and determinants of SARS-CoV-2 infection in selected communities in Zambia by examining the available updated population-based surveys in addition to the Hospital-surveillance data from selected communities in Zambia. The question is, how can we utilize the current trends and patterns of the SARS-CoV-2 infections to prompt rapid response preparedness and improved control and prevention strategies amongst high-risk groups? The study aimed also to identified the cluster patterns of this disease in the subpopulation, to further identify the risk populations and determinants like-wise. Despite the global decline in SARS CoV-2 incidences isolated cases continue being reported. WHO has not yet listed SARS CoV-2 as one of the eliminated or controlled pandemics thus the potential to emerge still exists especially with the massive mutations noticed!

## Materials and methods

### Study sites and design

The study was conducted in three (3) districts of Zambia; Ndola (Urban), Kafue (Peri-urban) and Lufwanyama (Rural). These sites were conveniently selected as they were SARS-CoV-2 Epi spots and places where low incidences were reported respectively. All eligible respondents at Ndola Central Hospital-Level 3 (Ndola), Lufwanyama district Hospital-Level 1 (Lufwanyama) and Kafue general hospital-Level 2 (Kafue) districts aged 15 to 65 years were selected and followed. Recruitment of participants and data collection was conducted from 7^th^ October, 2024 to 20^th^ December, 2024. All hospital cases between 11^th^ April, 2019 through to 15^th^ December, 2024 were eligible to be included. We performed an exploratory sequential mixed method study design using a multistage sampling technique. All SARS CoV-2 confirmed cases reported and recorded in the three study sites were eligible and all confirmed active SARS CoV-2 cases found during data collection. The mixed-method data collection approach is summarized in Table 1.

**Table 1:** Mixed-mode data collection and sampling to recruit participants in Ndola, Lufwanyama, Kafue district of Zambia October-December, 2024.

| Data Collection Mode | Target Number of Responses (n) | Geographic Reach | Sampling Approach | Further Notes |
| --- | --- | --- | --- | --- |
| Telephone surveys – tablet computers | 300 | Ndola, Kafue and Lufwanyama and residential areas / townships. | Snowball | Targeting people who do not have access to the internet and social media. Target was 100 per site |
| Face-to-face surveys – tablet computers | 30 | Ndola, Kafue and Lufwanyama residential areas / townships. | Simple random and Snowball | Targeting population survivals of COVID 19 have access to social media, internet, and telephone. Target was 10 per site |
| Hospital-based surveys | 480 | Ndola, Kafue and Lufwanyama residential areas / townships. | Simple random | Aim to review COVID-19 patient files from 3 hospital sites with target of 160 per site |
| Review SARS CoV-2 National and population-based surveys conducted in Zambia | 5 Publications | Lusaka and Copperbelt | Systematic review with search words: COVID 19, Zambia, Distribution, SARS CoV-2, Determinants | Aim to review observed trends of COVID-19 in these surveys |

### Sampling, sample size and data tools

The study employed mixed-mode sampling techniques for quantitative and qualitative data to be able to select files/records without any pre-selection as outlined in Table 1. Our calculated target sample was 363 respondents. The sample size was determined using the Krejcie and Morgan 1970s (21, 28) formula for a finite population.

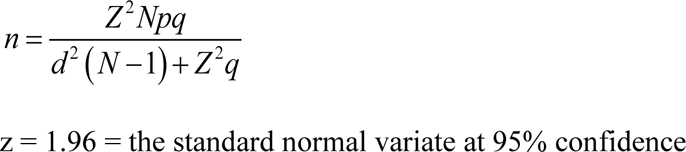

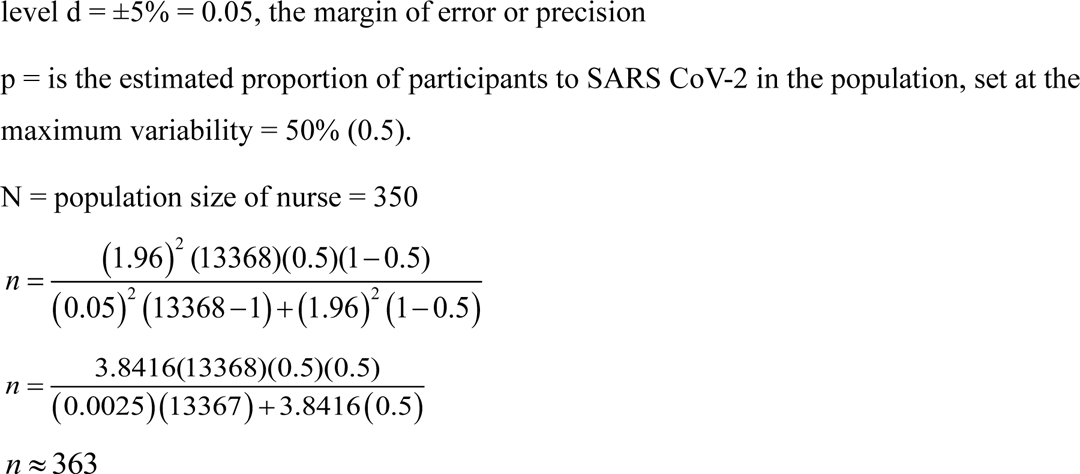

Assuming a 70% non-response rate, the study sampled 617 participants. Interest however was to collect as much data as possible and to follow all the active cases found during data collection. A summary of the data collection and sampling flow chart is in Table 1 and Fig 1. After obtaining written informed consent a detailed personal structured questionnaire was carried out with all eligible members in order to collect information on education, socio-demographic characteristics and risk behaviour, Knowledge, practices, vaccination status etc. All SARS CoV 2 confirmed active cases that were found were administered a questionnaire and an interview. Recent discharges (Only 1 actually) were tools administered on phone. In addition, a systematic review of selected point and period prevalence surveys conducted within Zambia in addition to National available data was conducted with the following search words, *“COVID 19, Zambia, Distribution, SARS CoV-2, Determinants.”*

**Figure 1:**
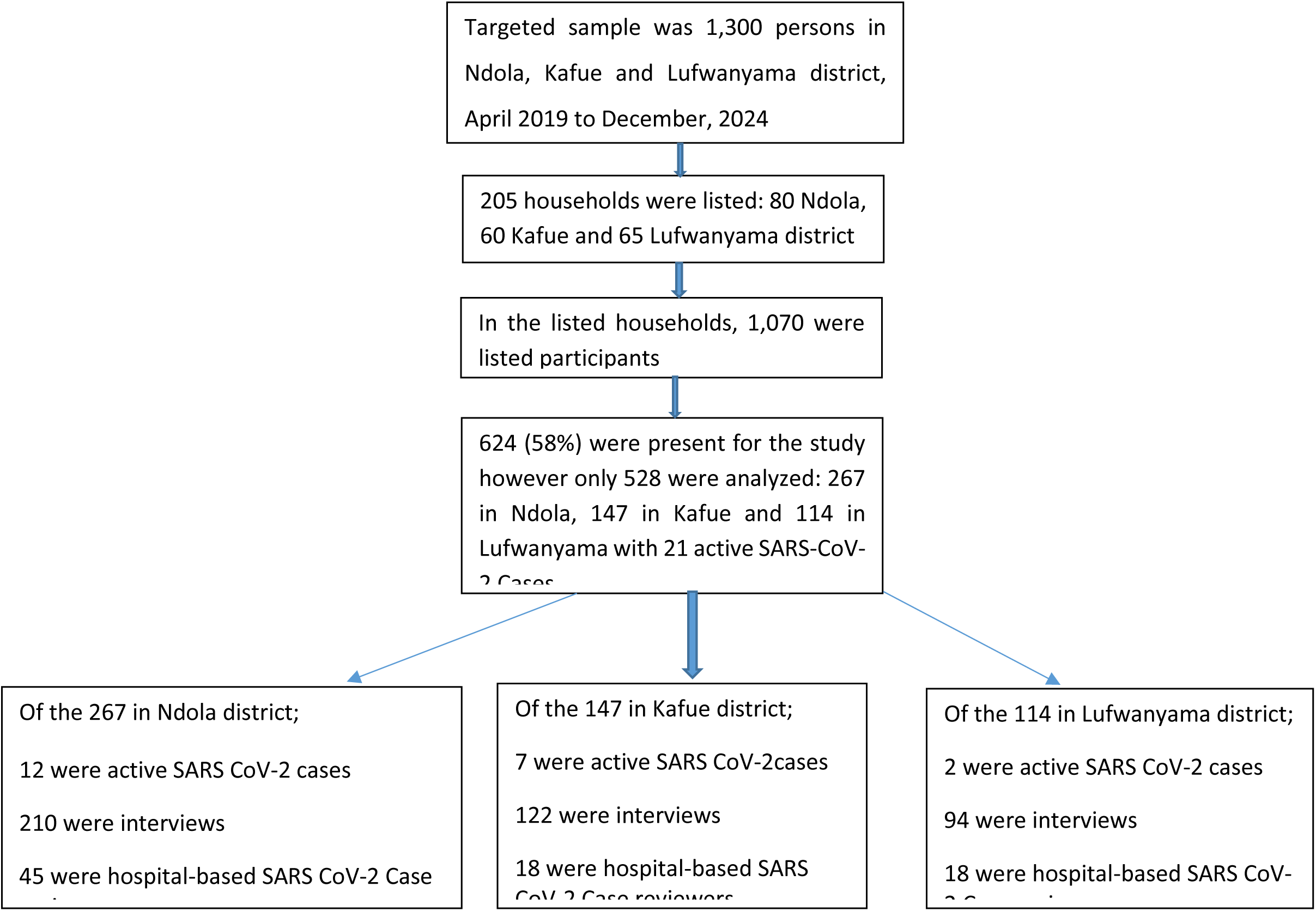
Flow chart of sampling process and indicative per site.

**Figure 2:**
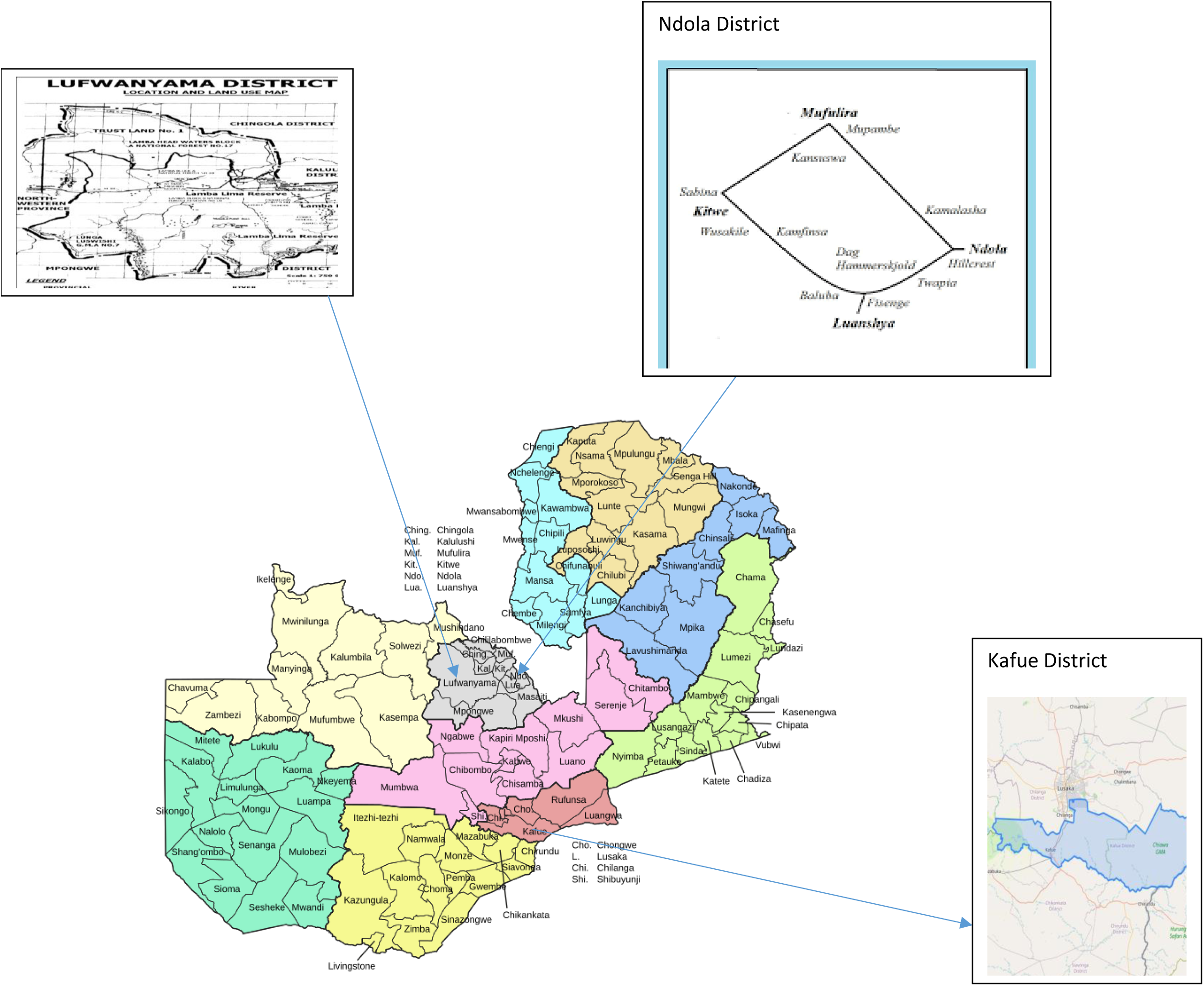
Map of Zambia with the three study sites in Copperbelt and Lusaka provinces, 2024.

**Figure 3:**
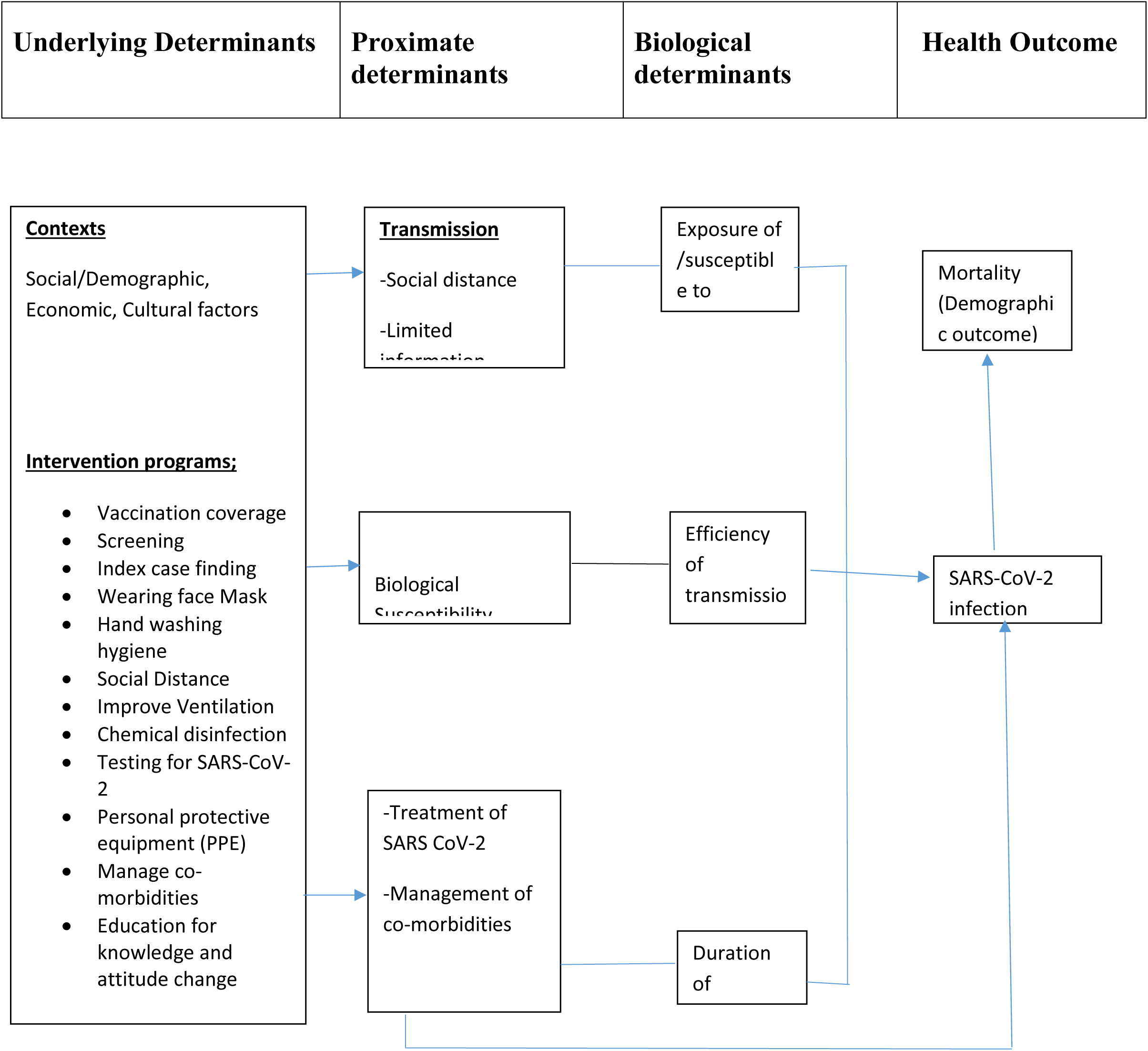
Proximate-determinant conceptual framework.

### Statistical analysis and ethical issues

Quantitative data was analyzed using Intercooled Stata version 12 (College Station, Texas, USA). Descriptive analysis has been presented as frequency tables and cross tabulations. The main statistical methods employed were the multivariate logistic regression, Pearson chi-square, and student’s t-test. Chi-square and Fisher exact tests were performed to test for differences in proportions of categorical variables between two or more groups. The level P < 0.05 was considered as the cutoff value or significance, exact and two-tailed. For qualitative analysis, thematic analysis was used to analyze data and correlate to the underlying determinants. Analysis used backward selection and excluded predictor variables highest p values singly until the final model containing only predictor variables with p < 0.05 was reached. To measure the strength of the association, both unadjusted and adjusted odds ratio were calculated. Our primary predictors of interest were age, sex, education, district, vaccination status, practices, experienced signs/symptoms, observed SARS CoV-2 guidelines. For all the face-to face interviews, written consent was obtained; for the hospital reviews verbal authorization was obtained from institution heads and for some telephone interviews verbal recorded consent was obtained witnessed by a second data collector after the participant was explained too. The study obtained consent from parents for 5 minors aged between 15-17. We obtained ethics approval from Chreso University Ethics Review Committee (Ref # 2254-09-2024) and National Health Research Authority (Ref # 2687/11/09/2025).

## RESULTS

### Participation and demographic characteristics

Overall (n= 624) respondents participated in this mixed-method study conducted in three different districts in Zambia; Ndola (267), Lufwanyama (210) and Kafue (147) with overall causes for non-participation been; refusal (27%), work-related (29%), absence (39%) and others (5%). The overall response rate for completing the interview was 78.9% with all the active cases having complete information. 15.4% (96/624) were not tested as their never had or found any record of a positive or negative SARS-CoV-2 test result either in their records or confirmation by interview thus were excluded in the analysis. Further, their case managements were not as SARS CoV-2 cases. The total number of cases which had tested or had confirmed record of SARS CoV-2 test conducted was 528. Details of participation in the study is summarized in Table 1 and Table 2.

**Table 2:** SARS CoV-2 sero status on data collection tools per district, 2024 (n=528)

| SARS CoV-2 sero status | Test status | Ndola (267) | Kafue (147) | Lufwanyama (114) | Total |
| --- | --- | --- | --- | --- | --- |
|  |  | Count (%) | Count (%) | Count (%) |  |
| Active cases found | Confirmed positive cases | 12 (4.5) | 7 (4.8) | 2(1.7) | 21 |
| Hospital-based reviews | Positive test result* | 2(0.7) | 1 (0.7) | 4 (3.5) | 7 |
|  | Negative test/record | 43(16.1) | 17(11.6) | 14 (12.3) | 74 |
| Interviews | Confirmed positive test | 6 (2.2) | 7 (4.7) | 8 (7.1) | 21 |
|  | No confirmation | 204(76.4) | 115 (78.2) | 86 (75.4) | 405 |
| Total Positives with confirmation/records |  | 20 (7.5) | 15 (10.2) | 14 (12.3) | 49<sup>1</sup> |
| Total Negatives with confirmation/records |  | 247(92.5) | 132 (89.7) | 100 (87.7) | 479 |
| Total |  | 267 | 147 | 114 | 528 |

\*Few showed no positive test result in records yet were managed as SARS CoV-2 cases (Symptomatically)
<sup>1</sup>Total number of all SARS CoV-2 positive cases

The mean age was 32 (SD: 10.3), there were more males 271 (51.3%) than females 257 (48.7%) in the study. In terms of marital status, the majority of those interviewed were (49.5%) married/cohabiting-Table 3. 80% listed home and work (67%) to be places where they had spent over 8 hours of their time respectively and 48.3% (255/528) had done self-diagnosis at home. 98.3% (519/528) believed that close contact with infected individuals can lead to infection, with only 2.3% having no knowledge of SARS CoV-2 transmission. Of 528, forty-nine (49) 9.3% had tested positive in all the study sites with 57.1% (28/49) of those having a positive antibody/nasal swab results found in their records and (21/49) having a confirmed positive SARS test done. Among the positives, 42.9% (21/49) were active cases found during data collection distributed as follows; 12 in Ndola, 7 in Kafue and 2 in Lufwanyama (Table 2). Of the 49 who tested positive, 27.4% assessed their self-risk as no chance, 18.7% very small chance and 53.9% as medium chance of contracting SARS-CoV-2, however there was no significant difference between the respective study sites (x^2^=3.2. p value 0.123).

**Table 3:**
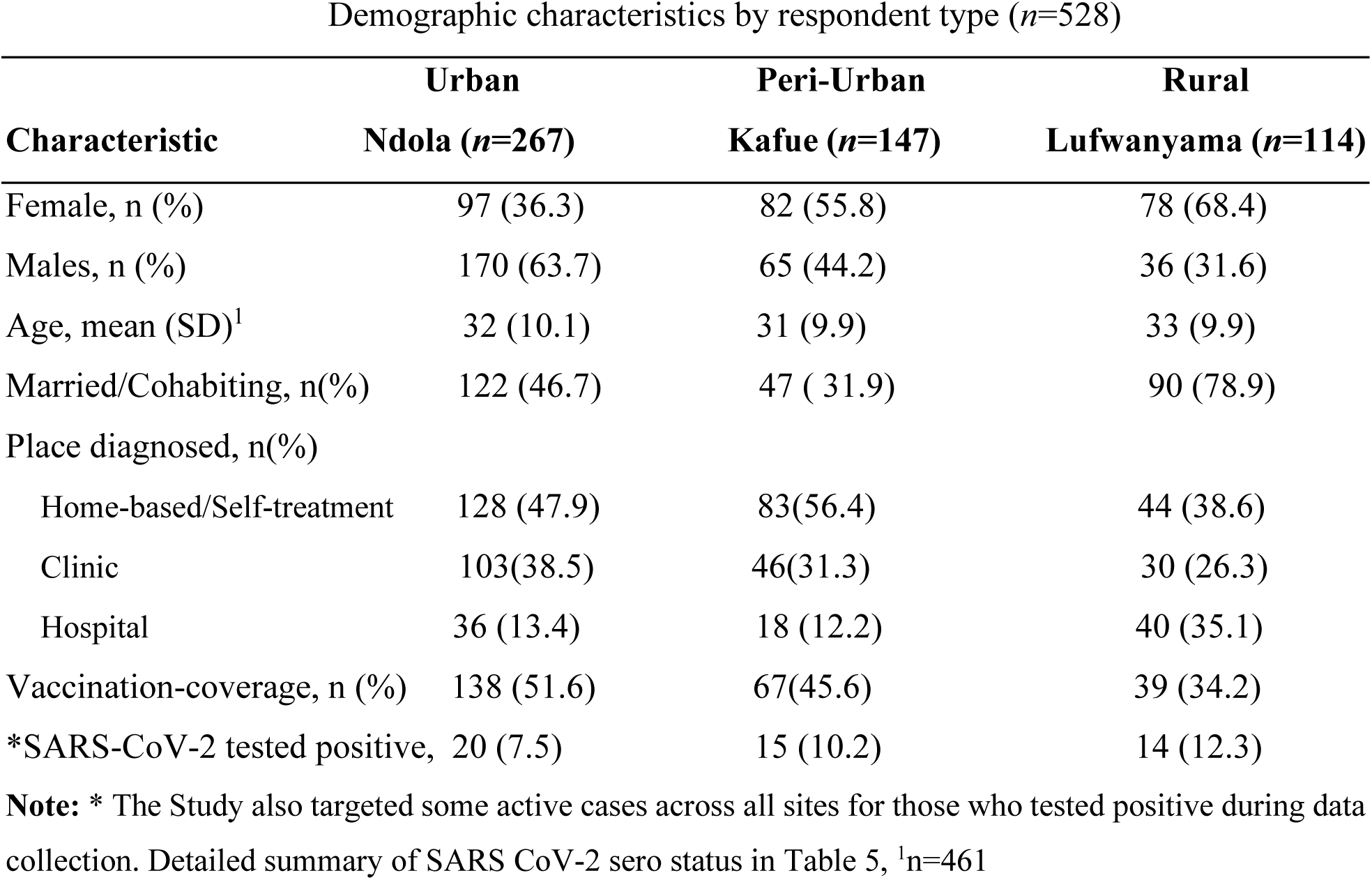
Social-demographic characteristics by site and respondent variable of available participants, 2024.

### Statistical associations, thematic and tests analysis

#### Underlaying determinants

In their initial explanations when asked what they knew about SARS-CoV-2, on average across study sites 492 (93.2%) discussed it as a disease caused by a virus and affects the respiratory system. Some differences in knowledge levels of knowing symptoms were observed among the respondents in Ndola and Lufwanyama district, otherwise most of them 487 (92.2%) did indicate that they knew the symptoms of SARS-CoV-2. No significant association was observed between SARS CoV-2 and education level (cOR 1.23 95%CI 0.79 1.88, p=0.354). Only 7.8% of the respondents indicated that they had no knowledge of how SARS-CoV-2 was transmitted.

Most respondents trusted information about SARS-CoV-2 from health care providers or Health centers (47.2%), radio or other social media platforms (32.5%) and 20.3% trusted information from Ministry of Health website. On the vaccination status, Ndola district had the highest vaccination coverage with over 51% of respondents having received their second doses with Lufwanyama having the lowest (34%). This however had no statistically significant difference among districts in SARS CoV-2 transmission (cOR 0.75 95% CI 0.49 1.17, p=0.208). Further, observing SARS CoV-2 prevention guidelines which included; Wearing face mask, hand washing hygiene practices, observing social distance was associated with reduced likelihood of infection (cOR 0.69 95% CI 0.45 0.88 p=0.012).

SARS CoV-2 infection distribution by person, place and time in the subgroups was categorized. It was observed that in older patients, SARS CoV-2 prevalence increase was more concentrated in males with comorbidities (cOR 2.12 95%CI 1.16 3.88). For example, in 2020, Mulenga *et al* (27) reported a pooled prevalence measure which increased with age. We found a significant association between SARS CoV-2 infection and sex (cOR 0.52 95% CI 0.28 0.95, p=0.034). Males were more likely to have SARS CoV-2 infection unadjusted for other variables. This study found that persons who spent more hours in high-risk places had great odds of infection. Other potential risk factors such as travel history, work hours interactions, chemical disinfection, ventilation and usual forms of transportation were not followed in this study.

Table 4 shows the results between participants social-demographic characteristics and SARS CoV-2 infection determined by using the Pearson’s chi square test. It can be observed that female had more positive tests than males, but there was not significant (P=0.112). The study also showed that there was no significant difference in positivity of those tests done in Ndola and Lufwanyama (x^2^=3.4, P=0.132). Most of the other social demographic characteristics such as age, marital status remined insignificant except level of education and the vaccination status/coverage between Ndola (Urban) and Lufwanyama (Rural), which was significant (P=0.006) but still remained insignificant between Ndola and Kafue (Peri-Urban) District (p=0.213).

**Table 4:**
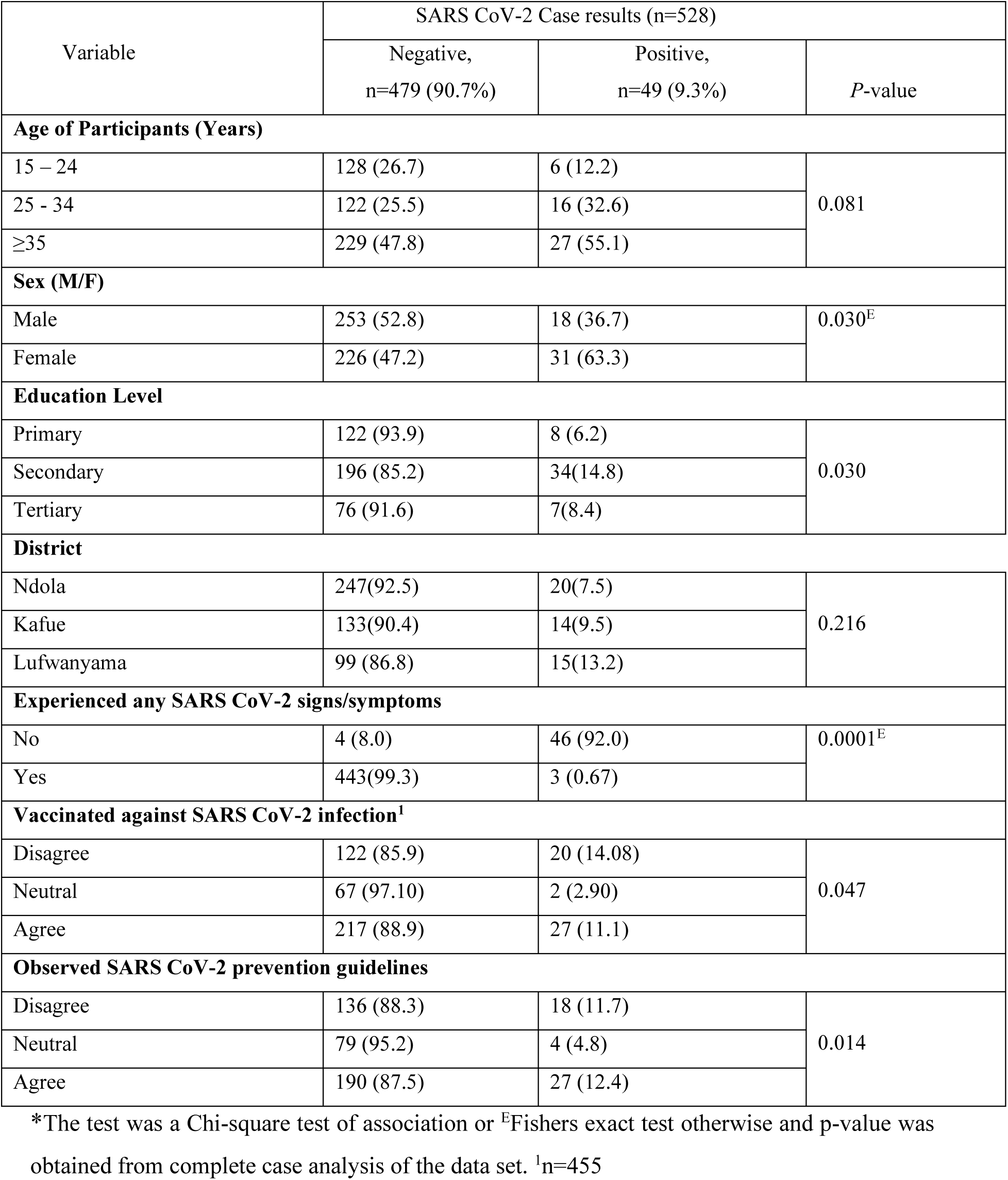
A summary of the cross tabulation of the social-demographic predictors of SARS CoV-2 sero status.

| Variable | SARS CoV-2 Case results (n=528) |  |  |
| --- | --- | --- | --- |
|  | Negative,<br>n=479 (90.7%) | Positive,<br>n=49 (9.3%) | P-value |
| Age of Participants (Years) |  |  |  |
| 15 – 24 | 128 (26.7) | 6 (12.2) | 0.081 |
| 25 - 34 | 122 (25.5) | 16 (32.6) |  |
| ≥35 | 229 (47.8) | 27 (55.1) |  |
| Sex (M/F) |  |  |  |
| Male | 253 (52.8) | 18 (36.7) | 0.030<sup>E</sup> |
| Female | 226 (47.2) | 31 (63.3) |  |
| Education Level |  |  |  |
| Primary | 122 (93.9) | 8 (6.2) | 0.030 |
| Secondary | 196 (85.2) | 34(14.8) |  |
| Tertiary | 76 (91.6) | 7(8.4) |  |
| District |  |  |  |
| Ndola | 247(92.5) | 20(7.5) | 0.216 |
| Kafue | 133(90.4) | 14(9.5) |  |
| Lufwanyama | 99 (86.8) | 15(13.2) |  |
| Experienced any SARS CoV-2 signs/symptoms |  |  |  |
| No | 4 (8.0) | 46 (92.0) | 0.0001<sup>E</sup> |
| Yes | 443(99.3) | 3 (0.67) |  |
| Vaccinated against SARS CoV-2 infection<sup>1</sup> |  |  |  |
| Disagree | 122 (85.9) | 20 (14.08) | 0.047 |
| Neutral | 67 (97.10) | 2 (2.90) |  |
| Agree | 217 (88.9) | 27 (11.1) |  |
| Observed SARS CoV-2 prevention guidelines |  |  |  |
| Disagree | 136 (88.3) | 18 (11.7) | 0.014 |
| Neutral | 79 (95.2) | 4 (4.8) |  |
| Agree | 190 (87.5) | 27 (12.4) |  |

\*The test was a Chi-square test of association or <sup>E</sup>Fishers exact test otherwise and p-value was obtained from complete case analysis of the data set. <sup>1</sup>n=455

#### Theme 1: Perspective on SARS CoV-2

In this theme, we present the findings on the perception of participants to the infection, responses to vaccination, knowledge on the disease preventions and/or signs together with awareness about the infection. All the 21 active cases interviewed demonstrate awareness to the signs and symptoms of SARS CoV-2 together with its prevention strategies. Some when asked why their got vaccinated, explained that it was the only way to get access to free movement. Others explained that, if you are not vaccinated you won’t be attending gathering. During discussions however, respondents noted that strict adherence to prevention guidelines by health providers, communities such as in churches, weddings, closure of schools and other social settings were key drivers to the increased vaccination rate especially in Kafue district

> “*At the health facility, when you come without a face-mask, they would send you back and tell you to buy a mask. [Only then] they will come and give you the drugs, that is when you’ll be attended to*.” –Participant, female, Lufwanyama district,

Yet another adds;

> “*Here in Kafue, the government people would go around with mega phones to encourage people to go for vaccination, so people can go back to their normal businesses. I want for vaccination, because I wanted to go back to the market*.”–Participant, female, Kafue district,”

In the subgroups, the cases were more concentrated in participants who spent most of their time in crowded places. On average 87.5% (462/528) spent their time at home and work. Interestingly, the persons who were observed to be at risk of severe cases of SARS CoV-2 infection were; elderly males above 49 years (aOR=7.16, 95%CI:0.12, 14.12, P<0.034), and those who had other co-morbidities or had past chronic respiratory infections (43%). Most of the participants in all the study sites had either a home treatment or self-diagnosis 255/528 (43.3%). The summary of the distribution of the levels of adherence to infection in the subpopulations agrees with some of the observations made during interviews of the perceived levels of adherence to preventive guidelines per location. (Table 5).

**Table 5:** Perceived levels of adherence in specific locations among communities by qualitative themes.

| Places with high adherence practices | Places with Lower adherence practices |
| --- | --- |
| ● School<br>● Churches<br>● Clinics | ● Markets<br>● Weddings<br>● Bars<br>● Within homes<br>● Soccer matches and work places |

The crude odds ratio (cOR) in the univariate analysis were performed (Table 9), which showed that participants who lived in Lufwanyama district had 1.4 times the odds of testing positive to SARS CoV-2 compared to participants in Kafue district (cOR=1.44, 95% CI:0.66, 3.12, p>0.356) though it was not statistically significant. On the other hand, participants who lived in Ndola district had 1.5 times the odds compared to participants who live in Lufwanyama (cOR=1.52, 95% CI:1.02, 7.62, p>0.231). Participants who had or their close associate had experienced SARS CoV-2 symptoms before had 2.8 times the odds of testing positive compared to those who never experienced the symptoms (cOR=0.0005, 95% CI:1.5, 6.92, p<0.0001). Participant sex (cOR=0.52, 95% CI:0.28, 0.95, p<0.034) and education level; Secondary (cOR=2.6, 95% CI:1.19, 5.90, p<0.018), tertiary (cOR=1.4, 95% CI:0.48, 4.03, p>0.528) compared to primary in Ndola and Lufwanyama district and other sites, had no statistically significant difference with testing positive for SARS CoV-2.

In the best fit model of the multivariable regression analysis highlighted in Table 9, stepwise regression model was performed by first running the logistic regression with all the predictor variables and then removing all the variables with the highest p-values one by one until we remained with the model that best explained the data-“best fit”. The predictor variables were; age, sex, education level, district, had symptoms or not, vaccination status, observed SARS CoV-2 prevention guidelines i.e., social distance and wearing face-mask, had co-morbidities, place spent over 8 hours per day. The best fit model had; age, vaccination status, had signs/symptoms, observing SARS CoV-2 prevention guidelines as significant explanatory (predictor) variables for sero-status of SARS CoV-2 infection in these subpopulations.

#### Theme 2: Factors influencing transmission of SARS CoV-2

Participants expressed different views and attitudes towards the preventive measures. Some explained that the measures were beneficial to the communities yet others wondered why they were being asked mixing with their friends and avoid social gatherings. Some further explained that at the beginning people didn’t pay attention to the prevention guidelines until their started noticing close relations dying. Of the participants who took part in the interviews, thematic analysis summary of 53% (16/30) of some participants said;

> “*After discharge, the doctor told me to get vaccinated and observe all the messages from Ministry of Health on prevention of SARS-CoV-2. He told me to avoid crowded places especially since I had TB*.” –Participant, female, Ndola district
>
> “*The places I noticed people were not following the guidelines were in funerals, weddings and beer halls. Because (they are) happy, dancing…. there don’t follow guidelines. At funerals they a lot of people, but those wearing masks are few*.” –Community member, female, Lufwanyama district

There was consensus among majority (62%) of participants concerning lack of adherence been one of the drivers to the increased transmission of the infection. When asked why some did not adhere to the prevention guidelines, most communities’ members believed the guidelines came from the whites and did not affect Africa. In all the study sites, there was free screening services for SARS-CoV-2 especially the elderly who had presented with respiratory infections and had other co-morbidities. Participants aged 49 years and above as well as having had experienced symptoms of SARS-CoV-2 had a significant relationship on them testing positive for SARS-CoV-2 (cOR=2.92, 95% CI:1.09, 7.28, p<0.047). Most respondent 255/528 (48.3%) had a home-based (Self-treatment) diagnosis after noticing some signs and symptoms of the infection. During the first and second wave, wearing a face mask in crowded places and observing social distance was made mandatory by government, however during the collection of this data, most persons did not observe this. In health care sittings such as clinics and hospital, only a few were observed adherering to this.

#### Proximate determinants

Most participants, 27/30 (90%) during interviews across all the study sites described coughing, sneezing, talking, and “exchanging air” as the main transmission modes of the disease. In addition, 67% agreed that not following recommended prevention methods increased the risk of transmission. Individuals who are highly susceptible to infections and had one or two significant underlying determinants increased the efficiency of transmission from one single cases.

Asymptomatic transmission (spread of infection by asymptomatic individuals) was not discussed in our present study across the three sites, though 7.9% had tested positive; 2.8% in Ndola and 5.1% in Kafue with none in Lufwanyama with no usual SARS CoV-2 signs and symptoms. This indicated that there could be some asymptomatic transmissions among populations. 477 (90.3%) did indicate that it was their personal responsibility to prevent themselves, their household and community against SARS CoV-2. Only 5.9% indicated that it was the government responsibility with 3.8% citing other agencies. In our current study, we did not discuss or determine the rate at which fomite, environmental factors, pH and radiation could impact on the transmission of SARS-CoV-2 among our populations.

The study however observed that most participants had home diagnosis or treatment as mentioned earlier. Of the 21 active cases across the three sites; records reviewed that there had not been vaccinated. 17 patients were above 24 years old and had other co-morbidities such as hypertension, diabetes mellitus, or history of chronic lung infection. The case fatality rate (CFR) as of period during date collection was 2/21 (9.5 per 100 or simply 9.5%). All the SARS CoV-2 positive cases from the records and active cases did indicate that most of them commenced treated on time with antibiotics and immune boosters. Other associated factors determined in this cohort is summarized in Table 6 and 7 respectively.

**Table 6:** Logistic regression adjusted and unadjusted Odds ratio with predictor variables, n=528.

| Predictor Variable | Unadjusted Odds ratio (95% CI) | P-Value | Adjusted Odds ratio (95% CI) | P-Value |
| --- | --- | --- | --- | --- |
| **Age, Years** |  |  |  |  |
| 15-34 | Ref |  | Ref |  |
| 35-48 | 2.13 (1.16,3.89) | 0.014 | 0.58 (0.032,10.5) | 0.719 |
| >49 | 0.13 (0.12,0.95) | 0.044 | 7.16 (0.124,14.12) | 0.034 |
| **Sex (M/F)** |  |  |  |  |
| Female | Ref |  | Ref |  |
| Male | 0.52 (0.28,0.95) | 0.034 | 3.68 (0.27,50.5) | 0.329 |
| **Marital Status** |  |  |  |  |
| Married/Cohabiting | Ref |  | Ref |  |
| Single | 3.3 (1.67,6.44) | 0.001 | 2.27 (0.14,36.08) | 0.561 |
| **Education Level** |  |  |  |  |
| Primary | Ref |  | Ref |  |
| Secondary | 2.6 (1.19,5.90) | 0.018 | 3.28 (0.92,3.23) | 0.986 |
| Tertiary | 1.4 (0.48,4.03) | 0.528 | 1.10 (0.36,2.28) | 0.987 |
| **District** |  |  |  |  |
| Kafue | Ref |  | Ref |  |
| Lufwanyama | 1.44 (0.66,3.12) | 0.356 | 1.44 (0.48,218) | 0.987 |
| Ndola | 0.77 (0.34,1.57) | 0.472 | 0.71 (0.23,12.32) | 0.988 |
| **Experienced any SARS CoV-2 Signs/Symptoms** |  |  |  |  |
| No | Ref |  | Ref |  |
| Yes | 0.005 (0.0001,0.002) | 0.0001 | 3.51 (1.32,5.46) | 0.946 |
| **Vaccinated against SARS CoV-2** |  |  |  |  |
| Agree | Ref |  | Ref |  |
| Disagree | 1.31 (0.71,2.44) | 0.383 | 4.20 (0.48,3.39) | 0.985 |
| Neutral | 0.24 (0.06,1.04) | 0.051 | 11.58 (0.47,285.1) | 0.134 |
| **Observed SARS CoV-2 prevention guidelines** |  |  |  |  |
| Agree | Ref |  | Ref |  |
| Disagree | 0.93 (0.49,1.76) | 0.827 | 2.37 (1.65,34.17) | 0.0123 |
| Neutral | 0.35 (0.12,1.05) | 0.067 | 7.4 (1.76,42.14) | 0.988 |

**Table 7:** Multivariable logistics regression.

| Variable, (Units) | Odds ratio | 95% Confidence Interval | P-Value |
| --- | --- | --- | --- |
| **Age, Years** |  |  |  |
| 15-34 | Ref | -1 |  |
| 35-48 | 1.02 | (0.16,6.62) | 0.983 |
| >49 | 2.92 | (1.09,7.23) | 0.047 |
| **Vaccination status** |  |  |  |
| Agree | Ref | -1 |  |
| Disagree | 2.17 | (0.18, 26.64) | 0.546 |
| Neutral | 14.87 | (0.97,228.63) | 0.053 |
| **Experienced SARS CoV-2 symptoms** |  |  |  |
| No | Ref |  |  |
| Yes | 0.0016 | (0.0013,0.0019) | 0.0001 |
| **Observed SARS CoV-2 prevention guidelines** |  |  |  |
| Disagree | Ref | -1 |  |
| Neutral<br>(Disagree/Agree) | 0.61 | (0.21,2.16) | 0.356 |
| Agree | 0.42 | (0.16,1.17) | 0.051 |

#### Biological determinants

The duration of infectivity was not determined in the study but traces of the viral antibody copies were still being detected even after 48 hours post completion of medication on an antibody test-serology (IgM/IgG) 11/21(52%). This can further be researched on as patients who would have completed treatment could still transmit (shed) the virus. 418 (79.2%) of the respondents and participants were working class, who spent over six (6) hours in their work places. The increased odds of transmission in such similar environments cited in table 8 to SARS CoV-2 infection is significant. Of 49 who tested positive 22 (45%) had home diagnosis with 27/49 (55%) despite some been vaccinated still testing positive. This could increase their exposure and transmission risk to the SARVS-CoV-2. 487 (92%) were high risk populations by age, co-morbidities, the environmental exposure and not vaccinated.

Despite the majority being knowledgeable about the risk factors and transmission modes of SARS CoV-2, individual characteristics and social behavior were predisposing them to the infection. The probability of estimating true positives (sensitivity) was 89.9% with 99.2 % specificity. Both the positive and negative predictive value were 95.6% and 98.1% respectively. The ROC value was 0.997 indicating that classification of SARS CoV-2 was not due to chance. Biologically, participants with old age and had other comorbidities were observed to be more at risk of SARS CoV-2 sero positivity and adherence to the preventive guidelines is one important prevention strategy that had led to decreased transmission and possible control of the infection. The efficiency of transmission from one single contact was not determined in this study due to few active cases rending the analysis insignificant. In addition, contact with a single confirmed case was rarely reported in rural areas compared to urban. Among the 21 symptomatic active cases, the most common reported signs/symptoms were; Headache (32.4%), chills (21.2%), fever (23.9%) and cough (22.5%). Applying this to urban or rural setting potential risk, the risk of transmission was more significant in the urban settings (X^2^=6.2, P=0.022).

## DISCUSSION

The aim of the study was to determine spatial epidemiology and cluster predictions on prevention strategies of SARS-CoV cases in selected districts of Zambia by identifying factors and determinates to the SARS CoV-2 transmissions in the populations. The study found a high rate of SARS CoV-2 infection in Zambian households in both rural and urban communities with the majority of these infections being asymptomatic. The results have shown that the estimated prevalence of SARS CoV-2 infection in Ndola, Lufwanyama and Kafue were 7.5%, 12.3% and 10.2% respectively. This prevalence decline in Ndola from 14.6% reported by Llyod *et al* (2021) (28) was concentrated in the elderly and educated groups. Kafue and Lufwanyama prevalence rate were among a few prevalence reported as baseline. No significant differences (p=0.28, χ² test) in SARS-CoV-2 prevalence were identified across districts, although prevalence was higher among people who resided in rural areas than in urban areas during the first wave. The later, sensitization and adherence could explain the variation. The increased incidences cases could have accounted also for the high prevalence accounting for the duration of the disease in the population.

These findings are more closely similar to studies which were done in other districts of Zambia -Kabwe, Livingstone, Lusaka, Nakonde, Ndola, and Solwezi)(28) except they reported a higher prevalence in urban areas. The findings have further demonstrated a pooled prevalence for the combined active SARS CoV-2 cases at 3.7% with CFR 9.5%. The reduced SARS CoV-2 cases and gradually decline in case reporting has partially led to some cases misdiagnosed or misclassified (8, 29, 30) Other studies conducted in Zambia reported a variation of between 5 to 15.8% of the population being infected in similar periods especially in areas (27, 31, 32) where there was less adherence to SARS CoV-2 prevention guidelines. In addition, other studies conducted (6, 28, 33) showed a higher prevalence of 15.7%.

The high prevalence suggests a high transmission rate within the subpopulations from one single infection or multiple infections. Other similar studies in contrast, conducted in Solwezi and Kalumbila by Mumba reported a 2.6% infection rate for all active case (34) with additionally other studies finding a much higher prevalence in similar settings, within the same period of survey, with highest infection rates occurring in March 2020 (26.3%) and lowest in March 2023 (0.86%). They did not however, correlate them to biological, underlying factors to estimate transmission rate. Our study suggests that people are aware about the SARS CoV-2 infection and the possible prevention strategies put to control it.

SARS CoV-2 being a respiratory infection, the spread of the infection can be increased by lack of adherence to prevention guidelines. Despite the massive enforcement and sensitization of the communities on SARS CoV-2 risk factors and prevention strategies, there were still new cases observed. The factors that could account for these cases are a discussion of this study. In 2021, Chen *et al* (7) demonstrated that transmission did not depend on the index case age or presence of symptoms but more on the index case RNA viral load. Other studies (35, 36) have attributed the transmission rate to lack of knowledge, poor attitudes towards prevention guidelines, environment (15) (Khan *et al.* 2021) and other biological factors in risk populations. The transmission patterns noticed in our study could be highly attributed to age, non-vaccination and lack of adherence to prevention guidelines.

Our findings showed that 80% had the knowledge of the infection. These findings corroborate previous studies which showed that most people have sufficient knowledge about the infection. Although the social order of human life is gradually returned to normal following the pandemic, new confirmed cases continued to be recorded worldwide. The majority of these cases were sporadic due to environmental factors and low levels of adherence to SARS CoV-2 prevention guidelines. Interventions focusing on improving personalized risk perception, behavioral beliefs and altitude have been shown to increase adoption of and adherence to a target health behaviour (6, 37-39).

Our finding further suggests that local social networks where individuals spend most of their time (Average 8 hours), frequency and duration of contacts are an important risk factor for the transmission of the virus. Despite the government restricting social gathering, most people maintained their social circles through social functions such as bars, weddings, church sessions and funerals (Table 9). Studies conducted by (9, 12, 40, 41) supported this notion thus policies regulating social interactions could help reduce the transmission of SARS CoV-2 infection. The transmissions are highly noticed to be clustered around such social circles. Hence, in emergency preparedness and rapid responses, identification and cases indexing of such circles is critical in the control of similar respiratory infections and should be considered. In addition, continued surveillance and monitoring populations at higher risk for a particular disease can be helpful to predict future outbreaks and focus prevention activities in the areas where they are most needed. The potential risks for epidemics due to continued new variants reported requires extensive research capacity and infrastructure in genomic surveillance and combined efforts towards one health for all.

Most of the cases in our study had no positive test result in their records or any confirmation during interviews yet many were managed as a SARS CoV-2 case. This supports WHO concern that most suspects wouldn’t present with the well-defined signs and symptoms and often some waited for any signs/symptoms to visit a health facility. This could have contributed to the high observed home diagnosis or treatment (Ave.57%) observed in this study. Further studies could be conducted on the irrational use of antibiotics especially Azithromycin on home use to treat SARS CoV-2 in Zambia (42). In addition, because of lack of testing kits in most rural facilities, symptomatic treatment was observed even in well-established health institutions.

The proximate determinate has clearly demonstrated the links and factors that could increase the health outcome-SARS CoV-2 infections. Of significant in our model was; age, vaccination status as underlying factors; observation of SARS CoV-2 prevention guidelines as a proximate determinant; and having experienced SARS CoV-2 signs/symptoms as a biological determinant. Participants who were above 49 years old; were not vaccinated; and also, never observed the SARS CoV-2 prevention guidelines and showed some signs or symptoms of the disease were at high odds of developing the outcome.

This finding adds further evidence apart from those reported in older patients above 60 years (28, 29, 43) that male adults above 49 years had greater odds (2.92, p<0.047) of risk. Among the active cases, those with such scenarios who also had some comorbidities were highly affected by the infection. At all cost, such risk groups should avoid risk environments (35, 44) and adhere to the prevention guidelines. Most of the vaccinated individuals showed low risks to the infection or the disease and did not continue into complications. With the herd immunity reached in Zambia as announced by MoH, the passive prevention exists in the population. In addition, adherence to the prevention guidelines in high-risk environments such as hospitals, crowded environments should continue to lower the risk of transmission.

The use of face masks, social distancing and hand washing facilities, in short, observing the SARS CoV-2 prevention guidelines were crucial in the control of the epidemic. Majority of the communities did not follow these guidelines. Asked why they did not adhere to the guidelines, most attributed it to bad attitudes. These public health strategies were crucial in reducing the risk of exposure and transmission. This was highly noticed in Kafue, where the Ministry of Health and other supporting partners enforced adherence. Prior studies on SARS CoV-2 suggests that respiratory droplets are the primary mode of transmission for beta coronaviruses (18, 44-46) which often according to the chain of infection is the portal of entry to a susceptible host.

Thus, proper case management could effectively contribute to low transmission rates. Index cases could as well be treated especially in household where there were elderly people above 49 years. There is substantial evidence to suggest and support CDC and WHO guidelines (2, 5, 15, 19, 22) that biological determinants, such as exposure rate, susceptibility and underlying determinants like social distancing, face masking can effectively reduce the transmission of the virus. There is however not enough evidence on how regular use of mask could contribute or the type of mask to reduce this transmission.

Another interesting finding of our study is the impact of the vaccination coverage on the transmission or pattern of cases from these districts. Kafue district, because of the partial lock down that was instituted in 2020 and the prevention strategies conducted such as; Health promotion, education, surveillance and enforcement of the SARS CoV-2 prevention guidelines resulted in a good response of the public towards vaccination by MoH. The critical feature of the proximate determinant framework linkage proximate determinants with contextual factors and intervention programs. For example, increased vaccination coverage an underlying factor could hypothetically result in zero transmission of cases. A number of vaccines were administered to individuals most commonly; the Johnson & Johnson and AstraZeneca which were globally accepted (29, 47, 48). Once herd immunity was reached (70%) (49) as indicated by USAID/MoH the vaccination had the potential to curb the infection (50). On the ground it was noticed that the majority (80%) of the population were well informed (knowledgeable) about the risk-factors, transmission rate and signs/symptoms of SARS CoV-2. Like in Kafue district, this was mostly attributed to the effective intervention programs during the lock down. More research however, is needed to determine the effective threshold for held immunity to sustain the disease pattern and trends in the population.

The findings presented and discussed in this report raise also many policies and research related questions as stated early such as; Firstly, the high number of self-treatment noticed could have future impact on microbial resistance on certain drugs that are also used for other common ointments. Ministry of Health should develop processes to control irrational use of such drugs in the general population. Suggested is for such drugs not to be over the count prescribed but with doctor prescription. Secondly, SARS CoV-2 was noticed to be more severe in the elderly with comorbidities. Deliberate programs targeted on such populations should be scaled up to quickly respond to such epidemic. For instance, all elderly at risk could be vaccinated as mandatory. The disease pattern, mortalities and trends (22, 37) clustered more among such subpopulations.

Thirdly, respiratory infections have still remained among the 10 major causes of death in Zambia communicable diseases profile in the past 12 years (24) Ministry of Health need to deliberately include the surveillance of SARS CoV-2 in the DHMIS system and national health surveys to monitor and survey the disease in the population. In addition, proper isolation and treatment facilities need to be available especially at Level 2 health facilities. Isolation of active cases, IEC-health education and promotion activities together with the vaccination of high-risk groups has been the focus on the control, management and prevention of SARS CoV-2 infection in Zambia. Zambia adopted the forementioned activities based on WHO recommendations and other locally studies (2, 3, 6, 18, 28).

Between March 1, 2019 and Dec 31, 2024 our model estimated the number and factors that could have influenced transmission rates within the 3 districts inferring that only about 1-4% (One in 10) of SARS CoV-2 infections in the respective regions were reported. The disease was more clustered in areas where individuals spent over 8 hours of their time and less adherence to SARS CoV-2 prevention. Individuals who had close associates experience SARS CoV-2 symptoms where higher odds of infection (cOR=0.0016, 95% CI:0.0013, 0.0019, p<0.0001). To foster rapid emergency response, the district health offices should be equipped with sufficient training to identify these high-risk clusters. In terms of the distribution of the disease by place, time and person; it was observed that places such as markets, bars had low adherence practices; further Individuals above 49 years old and not vaccinated stood at high risk of SARS CoV-2 infection. Work places were also clustered areas, where most transmissions took place especially if SARS CoV-2 prevention guidelines were not followed.

The study did have some limitations. Firstly, the design and findings are mostly based on few participants views. We could not conduct focused group discussions but we did include the interviews to minimize this scenario. The study was conducted during a period when there were few active cases of the disease thus difficult to analyse spatial epidemiology of cases. The only active cases found at the time of data collection were 21 insignificant for any spatial data analysis or follow up. We further recommend that more studies could be conducted to understand the spatial distribution of the infections in the population.

Despite these limitations, our study had several strengths. Firstly, we did combine several data collection techniques to corelate both quantitative and qualitative data i.e., Followed active cases, did hospital reviews of records as well as telephone interview of previous cases of SARS CoV-2. Secondly, triangulation through our purposive selection of the study participants from both rural and urban settings with varied socioeconomic backgrounds was essential to control some confounders and minimize bias. Finally, our study does provide insights on the distribution of the infection between rural and urban and also the links that exists via the proximate determinant to guide control of the disease.

## CONCLUSION

Majority of population were knowledgeable about the causes, transmission risks and the SARS CoV-2 prevention guidelines. Future prevention strategies should focus on; (1) Increase to access to SARS CoV-2 prevention guidelines (2) restricting social gatherings and (3) improving access to emergency response within districts (4) multi-sectoral genomic surveillance approaches and (4) integration of the SARS Cov-2 national health surveys is vital. Failure to adhere to the stipulated SARS CoV-2 prevention guidelines accounted for most of the increases transmission rate among the subpopulations.

## Data Availability

Data will be available and accessible through the corresponding author.

## ACKNOWLEDGMENTS

We thank Dr. Peter Mulenga, James musonda, Dr. Oliver Phiri and James Malakata for the assistance with the data collection and review of patient’s files. My gratitude to Dr. Gershom Chongwe who positively and academically criticized the article. Thanks to Edina Mulusa and Nancy Mudenda, for there consistent commitment in providing support and encouragement to continue with my research work.

## REFERENCES

1. Bahl A, Van Baalen MN, Ortiz L, Chen NW, Todd C, Milad M, et al. Early predictors of in-hospital mortality in patients with COVID-19 in a large American cohort. Intern Emerg Med. 2020;15(8):1485–99.

2. WHO. World Health Organization-COVID-19 Epidemiological Update. Geneva, Switzerland.: World Health Organization 2025 17 January 2025 Contract No.: 17 January 2025.

3. Progress report on HIV,Viral hepatitis and Sexual Transmitted Infection: Hearing before the WHO report, World Health Organisation(2019).

4. Dicker RA, Coronado F, Koo D, Gibson R. Principles of Epidemiology in Public Health Practice. Third ed. CDC: CDC; 2010. 512 p.

5. Massinga Loembe M, Tshangela A, Salyer SJ, Varma JK, Ouma AEO, Nkengasong JN. COVID-19 in Africa: the spread and response. Nat Med. 2020;26(7):999–1003.

6. Cohen C, Kleynhans J, von Gottberg A, McMorrow ML, Wolter N, Bhiman JN, et al. SARS-CoV-2 incidence, transmission, and reinfection in a rural and an urban setting: results of the PHIRST-C cohort study, South Africa, 2020-21. Lancet Infect Dis. 2022;22(6):821-34.

7. Chen N, Zhou M, Dong X, Qu J, Gong F, Han Y, et al. Epidemiological and clinical characteristics of 99 cases of 2019 novel coronavirus pneumonia in Wuhan, China: a descriptive study. Lancet. 2020;395(10223):507-13.

8. Zeitouny S, Suda KJ, Mitsantisuk K, Law MR, Tadrous M. Mapping global trends in vaccine sales before and during the first wave of the COVID-19 pandemic: a cross-sectional time-series analysis. BMJ Glob Health. 2021;6(12).

9. Wilkinson E, Giovanetti M, Tegally H, San JE, Lessells R, Cuadros D, et al. A year of genomic surveillance reveals how the SARS-CoV-2 pandemic unfolded in Africa. Science. 2021;374(6566):423-31.

10. Umviligihozo G, Mupfumi L, Sonela N, Naicker D, Obuku EA, Koofhethile C, et al. Sub-Saharan Africa preparedness and response to the COVID-19 pandemic: A perspective of early career African scientists. Wellcome Open Res. 2020;5:163.

11. Song H, Cao X, Ye H, He L, Li G, Wan T, et al. Pattern of COVID-19 in Sichuan province, China: A descriptive epidemiological analysis. PLoS One. 2020;15(11):e0241470.

12. Simulundu E, Mupeta F, Chanda-Kapata P, Saasa N, Changula K, Muleya W, et al. First COVID-19 case in Zambia - Comparative phylogenomic analyses of SARS-CoV-2 detected in African countries. Int J Infect Dis. 2021;102:455–9.

13. Sialubanje C, Sitali DC, Mukumbuta N, Liyali L, Sumbwa PI, Kamboyi HK, et al. Perspectives on factors influencing transmission of COVID-19 in Zambia: a qualitative study of health workers and community members. BMJ Open. 2022;12(4):e057589.

14. Lazarus R, Baos S, Cappel-Porter H, Carson-Stevens A, Clout M, Culliford L, et al. Safety and immunogenicity of concomitant administration of COVID-19 vaccines (ChAdOx1 or BNT162b2) with seasonal influenza vaccines in adults in the UK (ComFluCOV): a multicentre, randomised, controlled, phase 4 trial. Lancet. 2021;398(10318):2277–87.

15. Khan I, Shah D, Shah SS. COVID-19 pandemic and its positive impacts on environment: an updated review. Int J Environ Sci Technol (Tehran). 2021;18(2):521–30.

16. Kuijper A, Rookmaker MB, Mudrikova T. [Renal adverse reactions of antiretroviral medication: proximal tubular dysfunction associated with tenofovir]. Ned Tijdschr Geneeskd. 2011;155:A2249.

17. Burke RM, Midgley CM, Dratch A, Fenstersheib M, Haupt T, Holshue M, et al. Active Monitoring of Persons Exposed to Patients with Confirmed COVID-19 - United States, January-February 2020. MMWR Morb Mortal Wkly Rep. 2020;69(9):245-6.

18. Cabore JW, Karamagi HC, Kipruto HK, Mungatu JK, Asamani JA, Droti B, et al. COVID-19 in the 47 countries of the WHO African region: a modelling analysis of past trends and future patterns. Lancet Glob Health. 2022;10(8):e1099–e114.

19. Abdullah M, Ahmad S, Owyed S, Abdel-Aty AH, Mahmoud EE, Shah K, et al. Mathematical analysis of COVID-19 via new mathematical model. Chaos Solitons Fractals. 2021;143:110585.

20. Michelo C, Sandoy IF, Fylkesnes K. Marked HIV prevalence declines in higher educated young people: evidence from population-based surveys (1995-2003) in Zambia. Aids. 2006;20(7):1031–8.

21. Calleja JM, Marum LH, Carcamo CP, Kaetano L, Muttunga J, Way A. Lessons learned in the conduct, validation, and interpretation of national population based HIV surveys. AIDS. 2005;19 Suppl 2:S9–S17.

22. Jesri N, Saghafipour A, Koohpaei A, Farzinnia B, Jooshin MK, Abolkheirian S, et al. Mapping and Spatial Pattern Analysis of COVID-19 in Central Iran Using the Local Indicators of Spatial Association (LISA). BMC public health. 2021;21(1):2227.

23. Fwoloshi S, Hines JZ, Barradas DT, Yingst S, Siwingwa M, Chirwa L, et al. Prevalence of Severe Acute Respiratory Syndrome Coronavirus 2 Among Healthcare Workers-Zambia, July 2020. Clinical infectious diseases: an official publication of the Infectious Diseases Society of America. 2021;73(6):e1321-e8.

24. MoH Z. Zambia Demograohic Health Survey, 2024. MoH, Zambia: Zambia Statistical Agency; 2024.

25. Afriyie Osei D, Masiye F, Tediosi F, Fink G. Purchasing for High-Quality Care Using National Health Insurance: Evidence from Zambia. Health policy and planning. 2023.

26. Khan MR, Nazir MA, Afzal S, Sohail J. Health financing and public financial management during the Covid-19 pandemic: Evidence from Pakistan as low-income country. Int J Health Plann Manage. 2023;38(3):847–72.

27. Chanda D, Minchella PA, Kampamba D, Itoh M, Hines JZ, Fwoloshi S, et al. COVID-19 Severity and COVID-19-Associated Deaths Among Hospitalized Patients with HIV Infection - Zambia, March-December 2020. MMWR Morb Mortal Wkly Rep. 2021;70(22):807-10.

28. Mulenga LB, Hines JZ, Fwoloshi S, Chirwa L, Siwingwa M, Yingst S, et al. Prevalence of SARS-CoV-2 in six districts in Zambia in July, 2020: a cross-sectional cluster sample survey. Lancet Glob Health. 2021;9(6):e773-e81.

29. Zhou L, Ayeh SK, Chidambaram V, Karakousis PC. Modes of transmission of SARS-CoV-2 and evidence for preventive behavioral interventions. BMC Infect Dis. 2021;21(1):496.

30. Pung R, Lin B, Maurer-Stroh S, Sirota FL, Mak TM, Octavia S, et al. Factors influencing SARS-CoV-2 transmission and outbreak control measures in densely populated settings. Sci Rep. 2021;11(1):15297.

31. Chanda L, Tembo E, Sinyange N, Kayeyi N, Musonda K, Chewe O, et al. Characteristics of cases and deaths arising from SARS-CoV-2 infection in Zambia: March 2020 to April 2021. Pan Afr Med J. 2023;45:155.

32. Chileshe M, Mulenga D, Mfune RL, Nyirenda TH, Mwanza J, Mukanga B, et al. Increased number of brought-in-dead cases with COVID-19: is it due to poor health-seeking behaviour among the Zambian population? Pan Afr Med J. 2020;37:136.

33. Chu VT, Freeman-Ponder B, Lindquist S, Spitters C, Kawakami V, Dyal JW, et al. Investigation and Serologic Follow-Up of Contacts of an Early Confirmed Case-Patient with COVID-19, Washington, USA. Emerg Infect Dis. 2020;26(8):1671–8.

34. Mumba TK, Merwe KV, Divall M, Mwangilwa K, Kayeyi N. Seroprevalence survey of SARS-CoV-2, community behaviors, and practices in Kansanshi and Kalumbila mining towns. Front Public Health. 2023;11:1103133.

35. Acosta AM, Mathis AL, Budnitz DS, Geller AI, Chai SJ, Alden NB, et al. COVID-19 Investigational Treatments in Use Among Hospitalized Patients Identified Through the US Coronavirus Disease 2019-Associated Hospitalization Surveillance Network, March 1-June 30, 2020. Open Forum Infect Dis. 2020;7(11):ofaa528.

36. Maleche-Obimbo E, Attia E, Were F, Jaoko W, Graham SM. Prevalence, clinical presentation and factors associated with chronic lung disease among children and adolescents living with HIV in Kenya. PloS one. 2023;18(8):e0289756.

37. Kaiser JL, Hamer DH, Juntunen A, Ngoma T, Fink G, Schueler J, et al. COVID-19 Knowledge and Prevention Behaviors in Rural Zambia: A Qualitative Application of the Information-Motivation-Behavioral Skills Model. Am J Trop Med Hyg. 2023;109(1):76–89.

38. Li X, Xia WY, Jiang F, Liu DY, Lei SQ, Xia ZY, et al. Review of the risk factors for SARS-CoV-2 transmission. World J Clin Cases. 2021;9(7):1499–512.

39. Quaife M, van Zandvoort K, Gimma A, Shah K, McCreesh N, Prem K, et al. The impact of COVID-19 control measures on social contacts and transmission in Kenyan informal settlements. BMC Med. 2020;18(1):316.

40. Moore G, Rickard H, Stevenson D, Aranega-Bou P, Pitman J, Crook A, et al. Detection of SARS-CoV-2 within the healthcare environment: a multi-centre study conducted during the first wave of the COVID-19 outbreak in England. J Hosp Infect. 2021;108:189–96.

41. Msomi N, Lessells R, Mlisana K, de Oliveira T. Africa: tackle HIV and COVID-19 together. Nature. 2021;600(7887):33-6.

42. Gonzalez-Martinez C, Kranzer K, McHugh G, Corbett EL, Mujuru H, Nicol MP, et al. Azithromycin versus placebo for the treatment of HIV-associated chronic lung disease in children and adolescents (BREATHE trial): study protocol for a randomised controlled trial. Trials. 2017;18(1):622.

43. Barbhaya D, Franco S, Gandhi K, Arya R, Neupane R, Foroughi N, et al. Characteristics and Outcomes of COVID-19 Infection from an Urban Ambulatory COVID-19 Clinic-Guidance for Outpatient Clinicians in Triaging Patients. J Prim Care Community Health. 2021;12:21501327211017016.

44. Azuma K, Yanagi U, Kagi N, Kim H, Ogata M, Hayashi M. Environmental factors involved in SARS-CoV-2 transmission: effect and role of indoor environmental quality in the strategy for COVID-19 infection control. Environ Health Prev Med. 2020;25(1):66.

45. Bayin Donar G, Aydan S. Association of COVID-19 with lifestyle behaviours and socio-economic variables in Turkey: An analysis of Google Trends. Int J Health Plann Manage. 2021.

46. Brooks-Pollock E, Christensen H, Trickey A, Hemani G, Nixon E, Thomas AC, et al. High COVID-19 transmission potential associated with re-opening universities can be mitigated with layered interventions. Nat Commun. 2021;12(1):5017.

47. Odhiambo JN, Dolan CB. Spatial and spatio-temporal epidemiological approaches to inform COVID-19 surveillance and control: a review protocol. Syst Rev. 2022;11(1):141.

48. Odhiambo JN, Dolan CB, Troup L, Rojas NP. Spatial and spatio-temporal epidemiological approaches to inform COVID-19 surveillance and control: a systematic review of statistical and modelling methods in Africa. BMJ Open. 2023;13(1):e067134.

49. Celebrating attaining 70% COVID-19 full vaccination coverage [press release]. 01 November 2022 2022.

50. Linnisa W. Remarks for Deputy Chief of Mission Linnisa Wahid: Zambia Achieves 70% COVID-19 Vaccination Announcement. US Mission Zambia. 2022(State House, Lusaka):2.

